# A Contact-Explicit Covid-19 Epidemic and Response Assessment Model

**DOI:** 10.1101/2020.07.16.20155812

**Authors:** Wayne M. Getz, Ludovica Luisa Vissat, Richard Salter

**Affiliations:** Dept. ESPM, UC Berkeley, CA 94720-3114, USA; School of Mathematical Sciences, University of KwaZulu-Natal, Durban 4000, South Africa; Numerus, 850 Iron Point Rd., Folsom, CA 95630, USA; Computer Science Dept., Oberlin College, Oberlin, Ohio, OH 44074, USA

**Keywords:** SIR, corona virus, Policy evaluation, South Africa, England

## Abstract

We formulate a refined SEIR epidemic model that explicitly includes a contact class C that either thwarts pathogen invasion and returns to the susceptible class S or progresses successively through latent, asymptomatic, and symptomatic classes L, A, and I. Individuals in both A and I may go directly to an immune class V, and in I to a dead class D. We extend this SCLAIV formulation by including a set of drivers that can be used to develop policy to manage current Covid-19 and similar type disease outbreaks. These drivers include surveillance, social distancing (rate and efficacy), social relaxation, quarantining (linked to contact tracing), patient treatment/isolation and vaccination processes, each of which can be represented by a non-negative constant or an s-shaped switching flow. The latter are defined in terms of onset and switching times, initial and final values, and abruptness of switching. We built a [Covid-19NMB-DASA](https://www.cs.oberlin.edu/∼rms/covid3/) web app to generate both deterministic and stochastic solutions to our SCLAIV and drivers model and use incidence and mortality data to provide both maximum-likelihood estimation (MLE) and Bayesian MCMC fitting of parameters. In the context of South African and English Covid-19 incidence data we demonstrate how to both identify and evaluate the role of drivers in ongoing outbreaks. In particular, we show that early social distancing in South Africa likely averted around 80,000 *observed* cases (actual number is double if only half the cases are observed) during the months of June and July. We also demonstrated that incidence rates in South Africa will increase to between a conservative estimate of 15 and 30 thousand *observed* cases per day (at a 50% surveillance level) by the end of August if stronger social distancing measures are not effected during July and August, 2020. On different a note, we show that comparably good local MLE fits of the English data using surveillance, social distancing and social relaxation drivers can represent very different kinds of outbreaks—one with close to 90% and another with under 8% immune individuals. This latter result provides a cautionary tale of why fitting SEIR-like models to incidence or prevalence data can be extremely problematic when not anchored by other critical measures, such as levels of immunity in the population. Our presentation illustrates how our SCLAIV formulation can be used to carry out forensic and scenario analyses of disease outbreaks such as Covid-19 in well defined regions.

## 1 Introduction

The Covid-19 pandemic, currently sweeping the globe, is causing extraordinary disruption to the global economy and ripping apart the social fabric of society [1, 2]. Rational planning of policies implemented to avoid overloading healthcare systems, minimize death rates, and mitigate damage to various sectors of local and global economies requires the best available epidemiological model to underpin needed policy scenario evaluations [3, 4]. These epidemic models must reliably forecast the impacts of different human behavioral, societal healthcare, and patient care drivers related to key epidemic outcomes [5]. These outcomes, assessed by fitting models to regional incidence and mortality rate time series, include forecasts on hospitalization rates [6], extent to which incidence and mortality curves may be flattened through social distancing, quarantining, and other mitigating measures, as well as a possible second wave of incidence and mortality as mitigating measures are relaxed [7].

All epidemic models, whether systems or agent-based, are built around an SEIR (susceptible, exposed, infectious, recovered/removed) disease-class transition framework [8], with modifications to this framework appropriate to the particular disease process at hand. The basic SEIR framework, however, is inadequate for addressing certain key questions relating to the current Covid-19 pandemic. In particular, SEIR models do not explicitly separate out the contact and probability-of transmission-per-contact processes associated with disease transmission, a component key to exploring the impact of contact tracing on the epidemic. Also, SEIR models do not explicitly include a behavioral dynamics component related to reducing contact (i.e., social distancing) as incidence increases during the initial phase of the epidemic. In the context of Covid-19, it appears important to include both symptomatic and asymptomatic infectious state components, because this dichotomy has been identified as of considerable relevance to managing Covid-19 outbreaks [9].

SEIR models can, of course, be elaborated to include age-structure, which is useful when age-related effects on disease severity and mortality are evident [10], as is the case for Covid-19 [11]. Additionally, it is often useful for policy analyses to include a healthcare worker component [12], because special measures are needed to keep these workers active, but safe. At some point, these additional population structures should be included in future models, though at this time we can still learn much from homogeneous SEIR models, provided we confine our analysis to well bounded local or regional outbreaks where, as a first cut, spatial structure can be ignored [13].

The model we present here is an explicit contact compartmental version of the standard SEIR model that also includes, as found in several other Covid-19 models, both asymptomatic (A) and symptomatic (I) infections stages [14], as well as divides the exposed (E) stage into contact (C) and latent (L) disease stages. The contact compartment represent individuals that have been exposed to the pathogen, some of who return to the susceptible stage S having thwarted the infection through physical, physiological, and innate immunological mechanisms. Others in C succumb to infection and go on to the latent disease stage L (i.e., stage E in SEIR models). This division of E into C and L stages makes the contact process explicit rather than implicit, thereby allowing the effects of contact tracing to be explicitly incorporated into the dynamic model. Also, as discussed elsewhere [15], we make explicit that individuals sometimes lumped together in class R in SEIR models, have either recovered with immune (i.e., naturally vaccinate) in class V or are dead in class D. Finally, we include a set of SCLAI shadow or response compartments to hold individuals impacted by the processes, or drivers, used to manage the outbreak, including social distancing and subsequent social relaxation, quarantining in response to contact tracing, isolating/treating infectious individuals, and rolling out vaccination programs once suitable vaccines become available (Fig. 1). In addition, our model makes explicit the role of surveillance and reduced transmission associated with individuals in the response compartments of our SCLAIV+response model.

**Figure 1:**
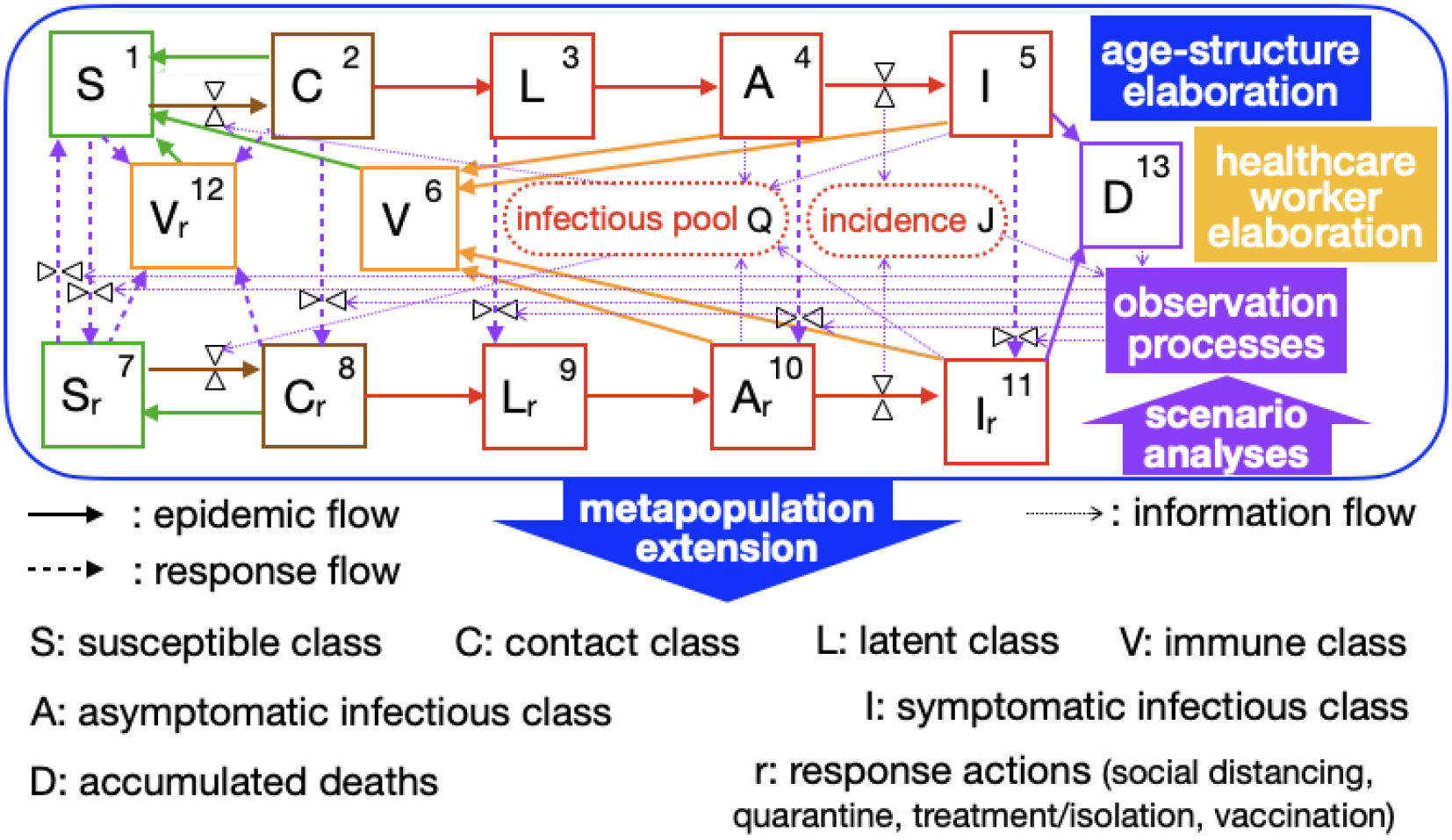
An epidemic and response flow diagram of a nuanced SEIR process that we refer to as a SCLAIV+response process. Within the infectious pool Q are two parameters that account for reduced transmission of asymptomatic compared with symptomatic individuals, as well as reduced transmission of individuals in response classes A_*r*_ and I_*r*_ compared with the original A and I classes. Within the incidence pool J, is a surveillance parameter that allows us to account for the fact that not all cases are observed. See text for more details on these parameters. The age-structure and healthcare worker elaborations are not developed in this paper, but are considerations for model refinement. The metapopulation extension can be used to incorporate spatial structure as discussed elsewhere [15]. Various scenario analyses can be undertaken, illustrations of which are considered in this paper using data from the current Covid-19 outbreaks in South Africa and in England, UK.

## 2 Model Structure

### 2.1 Contact Explicit Dynamic Model Formulation

In presenting the details of our SCLAIV+response model in this section, we emphasize that as with all basic SEIR models, populations are assumed to be well-mixed (i.e. any individual is equally likely to come in contact with any other individual in the population—so no relevent spatial structure) and are homogeneous from a behavioral and epidemiological characteristics point of view. In addition, we assume that natural birth and death processes, which—other than a disease induced process—cause population changes at time scales much slower than the epidemiological process under consideration and, hence, can be ignored.

Throughout this paper, we use the roman fonts S, C, L, A, I, V and D to name the disease classes defined below, while italic fonts *S, C, L, A, I, V* and *D* refer to the actual variables representing the number of individuals in these corresponding classes. We also introduce response classes SR, CR, LR, AR, IR, and VR, with variables *S*_*r*_, *C*_*r*_, *L*_*r*_, *A*_*r*_, *I*_*r*_, and *V*_*r*_ to respectively represent the number of individuals in each of these classes.

#### Disease classes

Our SCLAIV+response+D model consists of the following 13 variables (SCAIV is 6, response is 6, D is 1) with flow connections among classes depicted in Fig. 1.

1. *S*: number of susceptible individuals with pre-epidemic behavior
2. *C*: number of susceptible individuals who recently had contact with an infected individual
3. *L*: infected individuals in the latent/incubation stage (E class in SEIR models)
4. *A*: infectious individuals who are asymptomatic (they can next become either symptomatic or immune)
5. *I*: infectious individual who are symptomatic
6. *V*: recovered individuals with immunity (naturally vaccinated R class in SEIR model with no mortality)
7. *S*_r_: number of susceptible individuals with behavioral modifications in response to ongoing epidemic
8. *C*_r_: number of individuals in the *C* class who are quarantined
9. *L*_r_: exposed individuals who are quarantined
10. *A*_r_: infectious asymptomatic individuals who are quarantined
11. *I*_r_: infectious symptomatic individuals who are treated/hospitalized/isolated
12. *V*_r_: vaccinated individuals
13. *D*: individuals who have died from the disease (this is an absorbing state of the model and so, for clarity is listed last)

#### Embedded SCLAIV+response+D formulation

Our 13-variable SCLAIV+response+D model, apart from two flows, can be expressed in terms of a general “donor controlled” system where all per-capita flows out of compartments are either constant or time varying. The exceptions are the two contact processes in which individuals are transferred from S to C and from S_*r*_ to C_*r*_. These two flows, as we will see below, depend both on the value of the recipient compartment and the size of the active population as a whole (all individuals minus those that have died). Letting *γ*_*ij*_ represent the flow from compartment *j* to compartment *i* and using the numbering system of the variables listed above for our SCLAIV+response+D mode, we have 13 × 13 = 169 potential flows, but only 28 of the off-diagonal entries are non-zero, as indicated in the flow matrix Γ in Eq. 1 below (cf. Appendix A.1). Note that the last column of this matrix contains all zeros because *D* (*x*_13_, in the notation of Appendix A.1), is an absorbing compartment (no outflows, only inflows) and the remaining 12 diagonal elements are the negative sum of the rest of the columns in which they appear because the system is conservative (see Appendix A.1 for details).

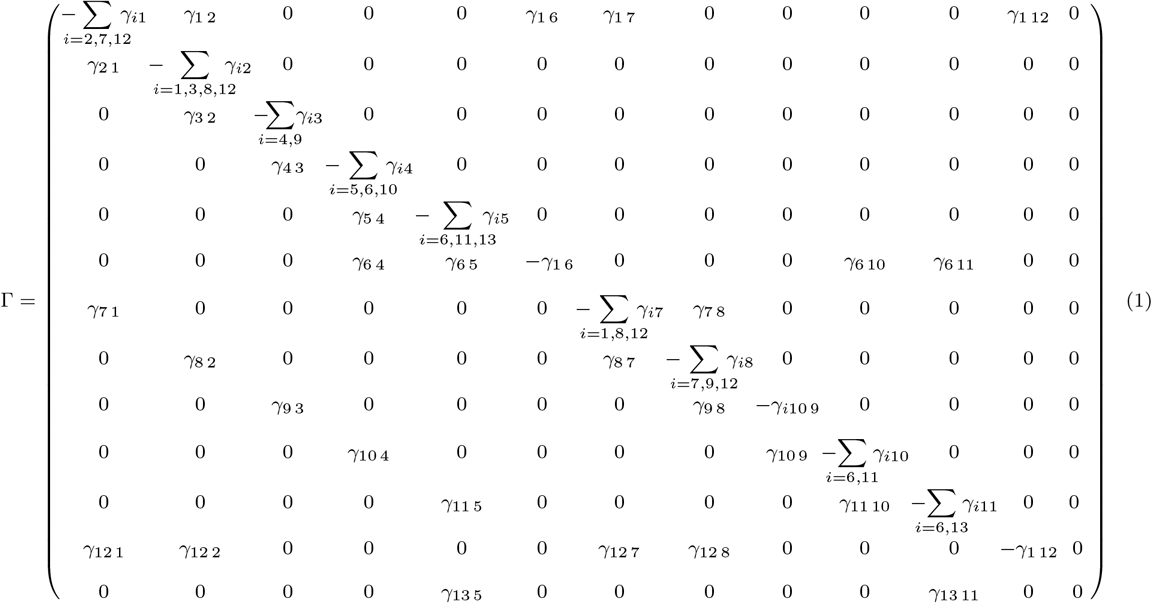

Our basic SCLAIV process is limited to the flows among the first 6 compartments (the top 6×6 submatrix of Γ defined in Eq. 1), which are assumed constant except for *γ*_2 1_, the contact process flow from S to C (i.e., *x*_1_ to *x*_2_ in the notation of Appendix A.1). This flow is assumed to depend on either direct contact with infectious individuals in disease class A and I (i.e., *x*_4_ and *x*_5_ in the notation of Appendix A.1)—or, more generally, with an infectious pool Q to which infectious individuals contribute, because some contacts are with viral-laden fomites rather than infectious individuals per se [16] (i.e. accounting for both direct and indirect modes of transmission, as discussed eslewhere [17, 18]). In a so-called frequency-dependent, rather than mass-action-dependent contact process, the intensity of the pool depends on the proportion of infectious individuals in the population, rather than the total number infectious individuals [19]. Similarly, in the broader the SCLAIV+response+D system, the flow *γ*_8 7_ from SR (*x*_7_) to CR (*x*_8_) is assumed to have this same frequency dependence, but is scaled down by a factor that accounts for reduced contact and, hence, transmission rates in social distancing individuals. The specifics of our formulation of the infectious pool Q, in terms of the number of individuals in classes A, I, A_r_, I_r_ and a total active (living) population variable *N* are provided by Eq. 12 below.

#### Contact rates and population size

In SEIR models, the processes of pathogen transmission and infection (i.e., successful pathogen invasion of a host once exposed to the pathogen) are concatenated and represented by a single expression *βϕ S*(*t*), *I*(*t*), *N* (*t*), where *β* is a force of transmission parameter and *ϕ* some appropriate functional form [20]. Specifically 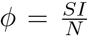 when assumed to be frequency dependent [19]. In our SCLAIV model, the parameter *β* is now separated out into the product of i) a force of contact rate parameter *κ* and ii) a proportion of individuals that succumb to (as opposed to thwarting) the infection once exposed to the pathogen (expression developed in Section 3).

In SEIR models, *N* (*t*) = *S*(*t*) + *I*(*t*) + *E*(*t*) + *R*(*t*) provided *R*(*t*) does not include individuals that have died from the disease. In our SCLAIV+response model, the comparable expression is

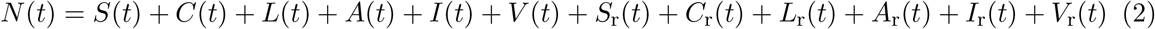

Before the infection begins, however, the size of the population at risk is often not known. Further, short of considering a closed population on an island, almost all populations “leak”: movement in and out occurs to some extent. Thus, when considering the size of the population at risk to Covid-19 in, say, London, New York, or Rio de Janeiro, a precise value to the size of the population at risk cannot be assigned. The solution to this problem is to normalize the analysis by standardizing the population size to a nominal value, say, *N*_nom_ = 10^5^ [5, 21] or *N*_nom_ = 10^7^ (as we do here for larger outbreaks), and then citing results per hundred thousand individuals (or per 10 million).

As long as the relative number of infected individuals in the population remains low the number of new cases remains largely independent of the size of the population at risk. Thus, epidemics start out generating new cases at similar rates in small towns and large cities, as long as the characteristic contact rates and epidemiological parameters in these different settings are similar. It is only once the epidemic gets going that towns run out of susceptibles much more rapidly than large cities and the epidemics in these contrasting situations begin to look quite different. In small towns the epidemic gets extinguished much earlier because it runs out of susceptibles, while in large cities it can continue to grown exponentially for a longer period of time. For this reason it is better to set *N*_nom_ = 10^7^ when modeling Covid-19 outbreaks in large cities or medium to large countries, provided this value is smaller than the true size of the population at risk, and the proportion of individuals in class V (a measure of the relative size of the epidemic) is less than, say, 10%.

### 2.2 Flows Rates

#### Basic SCLAIV process

An SEIR epidemiological process model, where R actually consists of both V (immune) and D (dead), is characterized by four parameters: the flows from S to E, E to I, I to V and a disease induced mortality flow from I to D. A fifth parameter is needed when the SEIR process is extended to the more general SEIRS formulation [8] (i.e., immunity is lost over time as individuals flow back from V to S). In a SCLAIV process, the splitting of E (exposed) into C (contact) and L (latent), and the addition of an A (asymptomatic) class now raises the number of parameters to the following eight:

1. the *contact rate* parameter *κ >* 0 scales the per-capita flow rate of susceptibles from class S to C in proportion to the per-capita intensity of the infectivity pool, where the reduced contribution to the pool by asymptomatic individuals, A, compared with symptomatic individuals, I, is scaled by an *infectivity reduction* parameter *ε* ∈ [0, 1). Specifically, the per-capita flow from S to C in the case of the SCLAIV model alone, for which the extant population is *N*(*t*) = *S*(*t*) + *C*(*t*) + *L*(*t*) + *A*(*t*) + *I*(*t*) + *V*(*t*), is

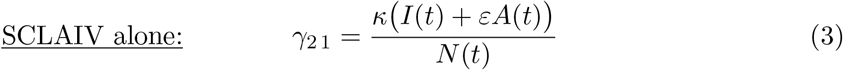 (Note: in an SEIR model *E*(*t*) = *C*(*t*) + *L*(*t*), *A*(*t*) = 0 and *V* (*t*) ≡ *R*(*t*))
2. the *succumb period* parameter *π*_suc_ ≥ 0 (i.e., pertaining to individuals who, after contact with the pathogen, succumb to infection), whose inverse is the per-capita flow rate

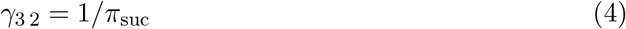

scales the flow from C to L
3. the *thwart period* parameter *π*_thw_ ≥ 0 whose inverse is the per-capita flow rate

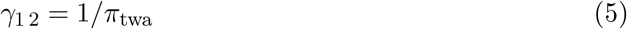

scales the flow from C back to S (i.e., those individuals who, after contact with the pathogen, thwart its invasion)
4. the *latent period* parameter *π*_lat_ ≥ 0, whose inverse is the per-capita flow rate

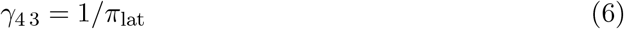

scales the flow from L to A
5. the *asymptomatic period* parameter *π*_asy_ *>* 0, whose inverse is the per-capita flow rate

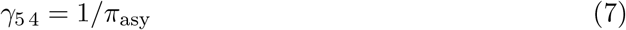

scales the flow from A to I.
6. the *infectious/recovery period* parameter *π*_rec_ *>* 0, whose inverse is the per-capita flow rate

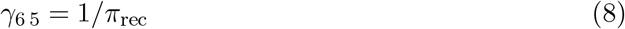

scales the flow from I to V. To keep things simple, we assume that the per-capita flow from A to V is scaled also by this same parameter i.e.,

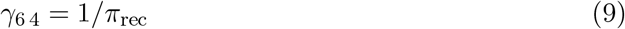

which implies that asymptomatic individuals can either play the role of presymptomatic individuals moving on to become symptomatic at a rate scaled by 1*/π*_asy_ or can recover from being asymptomatic at a rate scaled by 1*/π*_rec_
7. the *immune period* parameter *π*_imm_ ≥ 0, whose inverse is the per-capita flow rate

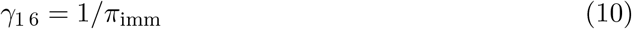

scales the flow from V back to S as immunity wanes over time
8. the *disease induced mortality rate* parameter *α* ≥ 0 is the per-capita flow rate from I to the proportion of D that represents in our SCLAIV+D process those dying from the disease: that is

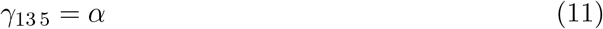

#### SCLAIV response process

The SCLAIV+response process can be divided along the following lines: 1.) a flow chain of individuals from SR to IR through CR, LR and AR and then onto V and D that parallels the SCLAIV+D process (Fig. 1); 2.) flows from basic disease classes to corresponding response classes (e.g., S to SR, C to CR etc.—see Fig. 1) through the implementation of drivers described in the next subsection used to develop policies for control and elimination of the outbreak. All but one of the first set of flows are governed by the same per-capita flow rates as in the basic SCLAIV process, the exception being the flow of individuals from SR to CR due to the reduced contact that individuals in SR have with the infectious pool Q, compared with the contact rates of S with Q.

The notion of a “virtual infectious pool” variable *Q* (tantamount to a measure of the amount of pathogen available for infecting susceptibles) allows us to represent the risk of making contact with the pathogen both directly (individual to individual) and indirectly (individual contacting a fomite) under the following two assumptions. First, four classes of individuals—A, AR, I, and IR— contribute to this infectious pool. Second, asymptomatic individuals (A, AR) shed only a proportion *ε* ∈ [0, 1) of pathogens that symptomatic individuals (I, IR) shed. Under these assumptions, a measure of the size/intensity of the infectious pool is

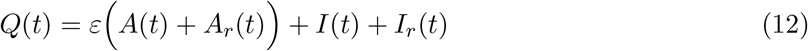

The per-capita rates at which susceptible individuals encounter this pool at time *t* is now assumed to be proportional to the of size of the pool *Q*(*t*) normalized by number of individuals *N* (*t*) (Eq. 2) available for contact. We also let *δ*_con_ ∈ [0, 1) represent the extent to which social distancing behavior reduces contact with infectious pool *Q*. In this case, equation 3 can be generalized to obtain the flow rates

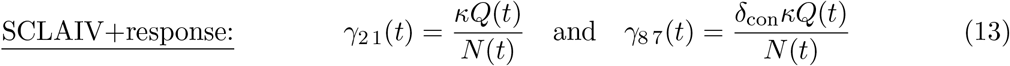

For the sake of completeness, the assumption that remaining response compartments are the same as in the basic SCLAIV process implies that

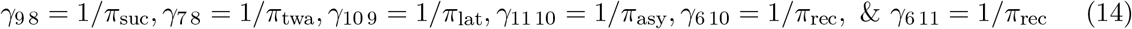

and, for simplicity, we also assume that

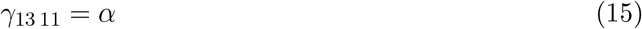

In reality, the mortality rates in compartments I and IR are bound to differ, but the change in mortality rate over time is, perhaps, better dealt with as a driver that leads to a reduction in mortality rates once the medical establishment has improved its protocols and therapeutics for treating individuals infected with the disease. Note, under the assumption that the surveillance and treatment/isolation drivers are activate from the start of the epidemic, during the course of an outbreak most individuals land up in compartment IR rather than I itself. On the other hand, individuals will not be shunted to compartments CR, LR and AR unless contact tracing along with quarantining is implemented.

### 2.3 Response Drivers

The response drivers, which are responsible for transfer individuals from the SCLAIV set of classes to the SCLAIV-response classes (Fig. 1) can either be specified to have some constant value or may be represented in terms of switching curves when time varying. Specifically, when represented in terms of switching curves, we assume that the drivers are 0 until they are initiated at some point in time, either at the start of the outbreak or sometime into the outbreak, and that they may either represent a gearing up process, as in surveillance, social distancing and quarantining, or a gearing down process, as the relaxation of social distancing and reductions in disease-induced mortality rates.

The classic curve for modeling a graduated switch from a value of 0 to 1 on the infinite line (−∞, ∞) is the sigmoidal or logistic curve with two parameters: the first specifies the switch point (i.e., the point of inflection on the curve) and the second specifies the slope of the curve at the switching point. This function involves the exponential that, when replaced with a power function, switches on [0, ∞). This new related function can be rescaled using two additional parameters and translated by a constant for the addition of one more parameter to have it turn on at some time *t*_0_ and switch between initial and final values at some switching time *t*_1*/*2_ (Fig. 2). Specifically, in the context of a particular driver named “x” we define a function that switches between two values *δ*_x 0_ and *δ*_x ∞_ on the interval the half-infinite interval [*t*_x 0_ ≥ 0, ∞] by the equation

**Figure 2:**
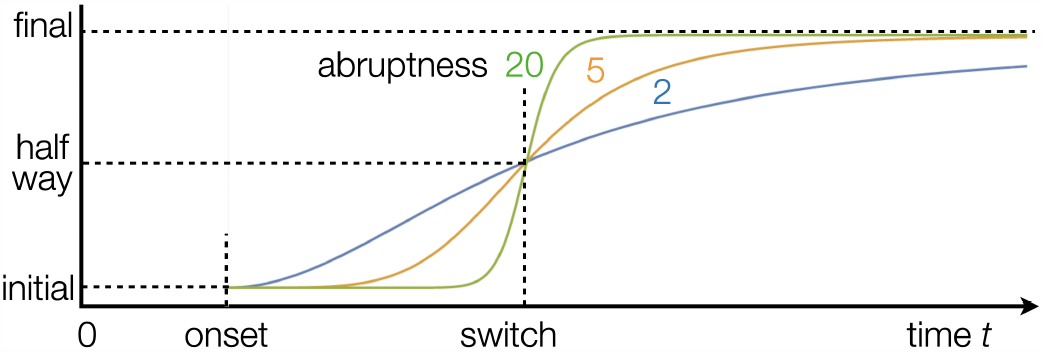
Switching curves (Eq. 16) used to represent the time-course of drivers included in our SCLAIV response process are 0 until onset time *t*_0_ at an initial value (*δ*_0_; dropping the “x” designator in Eq. 16). They then switch to a final value (*δ*_∞_) by passing through the “half-way” point at time *t*_1*/*2_ with an “abruptness” or steepness specified by the parameter *σ >* 1 (strict inequality ensures that the derivative of *δ*(*t*) is zero from the left at *t*_0_) and the switch becomes increasingly sharp to become a step-function as *σ* → ∞. If the initial value is larger than final value the switch is “off” rather than “on.”

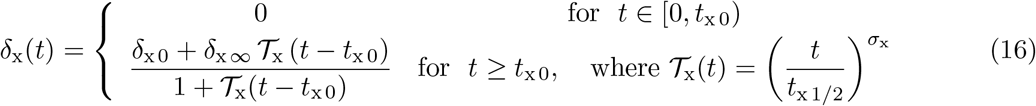

We note that for this driver, the time scaling function 𝒯_x_(*t*) implies that the switch is half-way complete at time *t* = *t*_x 1*/*2_ and that the abruptness of this switch (i.e. steepness at *t*_x 1*/*2_) is controlled by the value *σ*_x_ [22]: as *σ*_x_ → ∞ the switch approaches a step function that switches from *δ*_x 0_ to *δ*_x ∞_ at *t* = *t*_x 1*/*2_. Typically *σ*_x_ ∈ [2, 20] is sufficient to go from a gradual switch to an almost instantaneous switch (Fig. 2). Also, if *δ*_x 0_ *< δ*_x ∞_ then curve switches up (on) otherwise it switches down (off). Also, if we want to ensure that 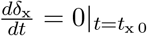, then we need to select *σ*_x_ *>* 1.

In the formulation of our model, we allow the following parameters/flow-rates to either be constant (usually when fitting initial conditions and contact and transmission rate parameters in the model to the initial outbreak phase) or to be represented as drivers that can be switched up or down as a means to managing an outbreak. When represented as switching functions, each of the drivers below follows Eq. 16 with x replaced by a three letter acronym as indicated.

**Surveillance response driver**, *δ*_sur_(*t*), determines the proportion of new cases identified in each time period through an equation presented below in the context of our discrete time analogue of the continuous time model that has the flow topology of equation 1.

**Social distancing driver**, *δ*_sod_(*t*) ∈ [0, 1), determines the per-capita flow rate from the susceptible class S to SR: i.e., *γ*_7 1_ = *δ*_sep_(*t*)

**Social relaxation driver**, *δ*_sor_(*t*), is used to set the flow rate *γ*_1 7_ from susceptible class SR back to S using the relationship *γ*_1 7_ = *δ*_rel_(*t*)

**Quarantine response driver**, *δ*_qua_(*t*), determines the per-capita flow rates *γ*_8 2_, *γ*_9 3_, and *γ*_10 4_ from classes C, L and A to CR, LR and AR respectively: i.e., *γ*_8 2_ = *γ*_9 3_ = *γ*_10 4_ = *δ*_qua_(*t*)

**Patient isolation/treatment driver**, *δ*_iso_(*t*) determines the per-capita flow rate of symptomatic individuals in class I to patient care/treatment class IR: i.e., *γ*_11 5_ = *δ*_iso_(*t*)

**Vaccination rate driver**, *δ*_vac_(*t*) determines the per-capita flow rate at which individuals are vaccinated, should on exist, during the course of the epidemic: i.e., *γ*_8 2_ = *δ*_vac_(*t*)

**Contact rate reduction driver**, *δ*_con_(*t*) ∈ [0, 1], is needed to compute *γ*_2 1_ using Eq. 13

Lastly, one can also express the disease-induced mortality rate *α* as a switching function, but we have not done so because we have not treated reductions to disease-induced mortality as a driver in this formulation. Rather, we view this reduction as driven by a much longer term therapeutics development process that, as yet, cannot be implemented on the time scale of the various drivers considered above.

## 3 Discrete Time Implementation

### 3.1 Competing Rates Formulation

Continuous time formulations of SEIR processes have notational advantages over discrete time formulations in terms of presentation and the development of theory, such the derivation of stability results, including in metapopulation situations [23], and the identification of backward bifurcation phenomena [24]. Discrete-time formulations, however, are more directly related to incidence and mortality data (reported daily in the case of Covid-19), simpler to numerically simulate, and more easily extended to stochastic settings [15, 25]. Of course, individual-based models provide the greatest flexibility of all when it comes to dealing with stochasticity—particularly in the context of following transmission chains [26] or dealing with all manner of heterogeneities [27] and proliferation of diversity in the pathogen itself [28].

When outbreaks are relatively large—on the order of tens of thousands or more individuals—and the focus is on developing policy for managing an outbreak rather than exploring questions relating to host and pathogen heterogeneity, then a discrete time systems formulation provides the most efficient way to undertake scenario analyses. The most consistent way to discretize a continuous systems model represented by the flow matrix Eq. 1 is to use a competing rates approach to obtain the analogous equations [15]. Specifically, the equations so obtained are

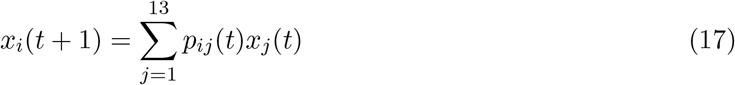

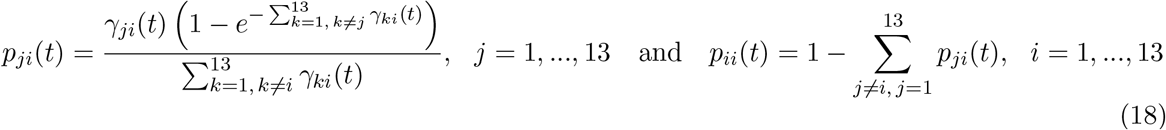

where the matrix *P* (*t*) with elements *p*_*ij*_(*t*), *i, j* = 1, …*n*, have the structure depicted in Fig. 3.

**Figure 3:**
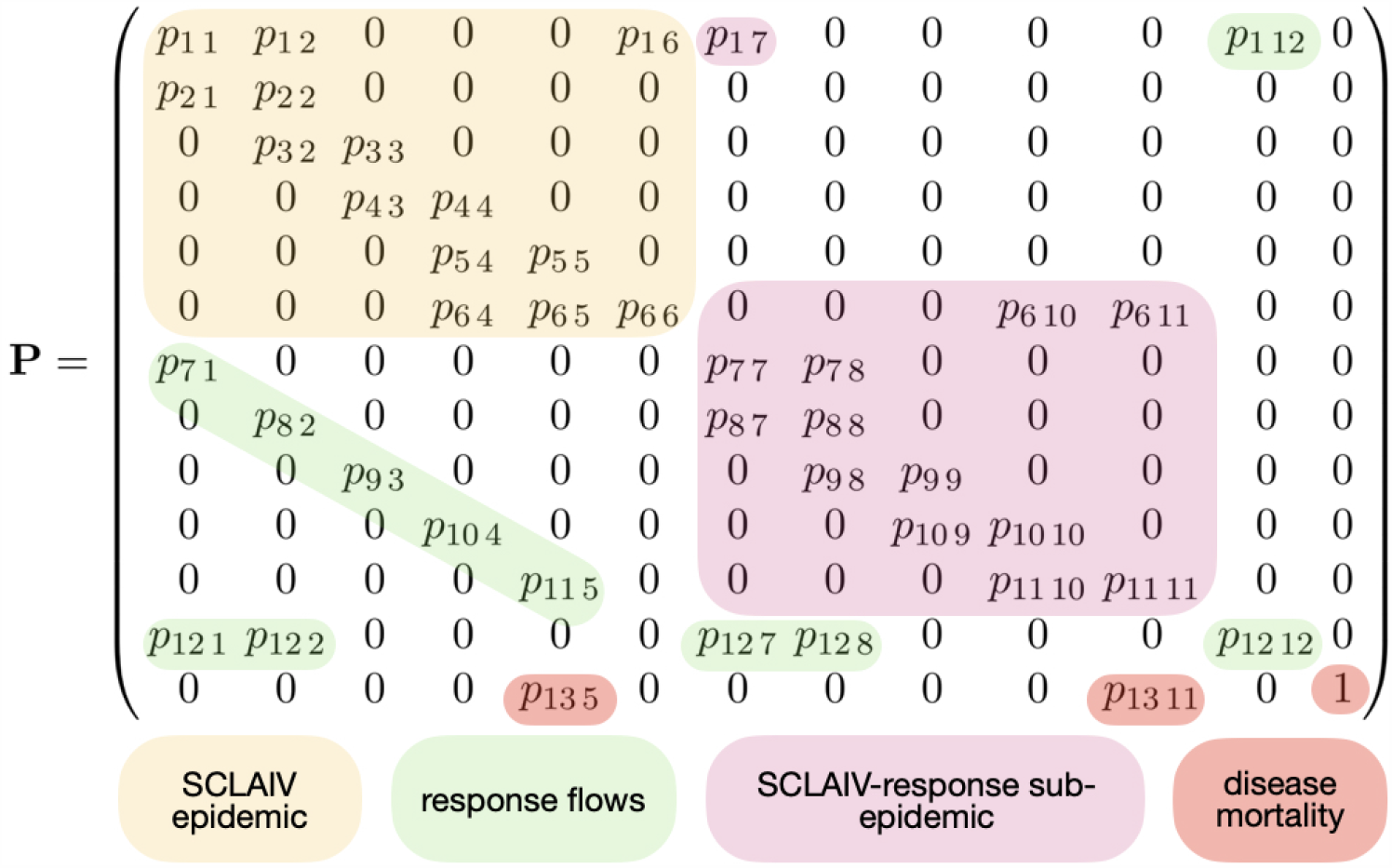
A depiction of the flow topology implied by the sparse structure of the stochastic matrix *P*, which can be used to carry out deterministic/stochastic simulations of the SCLAIV+response+D process when the elements *p*_*ij*_ in the matrix *P* are treated as proportions/probabilities of individuals transferring from disease class *j* into disease class *i* at time *t*. Color-coded blocks of elements pertain to transfers under the basic SCLAIV process (yellow) the parallel SCLAIV response process (pink), the SCLAIV response-driver processes (green) and the mortality process (red)

As mentioned earlier, the force of transmission parameter *β* in the SEIR formulation is defined to be the per-capita contact rate *κ* multiplied by the proportion/probability *p*_3 2_ of individuals succumbing to infection having made contact with the pathogen. Using Eqs. 4, 5 and 18 to express this proportion in terms of the periods *π*_thw_ and *π*_suc_, *β* can now be expressed as

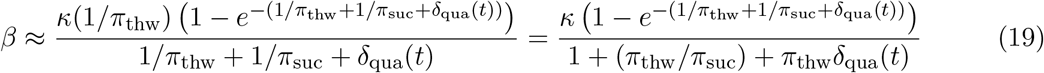

Note we use ≈ rather than strict equality because *β* and *κ* are rates in continuous time formulations while the proportion succumbing to infection is computed using a competing rates approximation that applies over one unit of time (i.e., we are mixing a continuous time process with a discrete time event). Thus the estimate *β* will depend on the units of time used in the formulation (e.g., days for relatively fast spreading diseases versus weeks or months for slower spreading diseases).

In addition, we note that these proportions in Eq. 18 are constants whenever the values *γ*_*ij*_ used to compute them are constant. If any *γ*_*ij*_(*t*) are time varying, either because they are computed from drivers or part of the transmission process (Eq. 13), then as the values change they are approximated by constants over each time interval [*t, t* + 1), but change from the start of one time interval to the next as the solution numerical unfolds (i.e., is iterated from *t* = 0 to final time *t* = *T* using the transition matrix *P* depicted in Fig. 3 <u>[15]).</u>

The conservative property of the matrix Γ, given by Eq. 1 (i.e., all column sums are zero) implies that all columns of the transition matrix *P* (*t*) (Fig. 3) sum to 1. We can either interpret the elements of the matrix *P* as proportions (deterministic formulation) or as probabilities. The latter applies in the case of stochastic simulations (i.e., the matrix *P* becomes a “stochastic matrix”) and each simulation is then one instatiation of a Markov chain process. Thus we can either use the matrix *P* to generate deterministic or stochastic simulations, as described in [15].

### 3.2 Model Fitting

Our basic SCLAIV formulation, under simplifying assumptions that we introduced along the way, requires values to be estimated for the following nine parameters introduced in Subsection 2.2: *κ* (force of contact), *ε* (reduced infectiousness of asymptomatics), *π*_suc_ (succumb period), *π*_thw_ (thwart period), *π*_lat_ (latent period), *π*_asy_ (asymptomatic period), *π*_rec_ (recovery from infection period), *π*_imm_ (immune period), and *α* (disease induced mortality rate). In addition, the 6 initial conditions *S*_0_, *C*_0_, *L*_0_, *A*_0_, *I*_0_, and *V*_0_ need to be set, assuming *D*_0_ = 0.

In fitting a basic SCLAIV process to a set of initial outbreak data, it may be reasonable to assume that all of the response drivers, apart from a background surveillance and isolation of the

sickest patients, are switched off. Surveillance needs to be operating at some initial level *δ*_sur 0_ (which either implies that *t*_sur 0_ = 0—see Eq. 16 or we can treat surveillance as a constant rather than a switching function) if any cases are to be observed. If we assume in our model that, initially, a proportion *δ*_sur 0_ ∈ [(0, 1] of new infectious cases are observed, then the number of new infectious cases *O*(*t*) observed during time period *t*, expressed in terms of the proportion *p*_5 4_ of individuals transferring from A to I, is given by

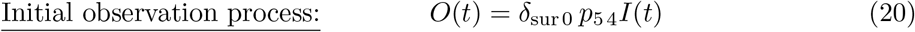

Once the full SCLAIV+response process is underway the observation process now includes the proportion *p*_11 10_ of individuals transferring from AR to IR as well. In this case the observation process, which now may depend on a surveillance driver *δ*_sur_(*t*) that is ramping up over time, is given under the assumption that the same surveillance level applies to class *I*_r_ (of course a different assumption can be made)

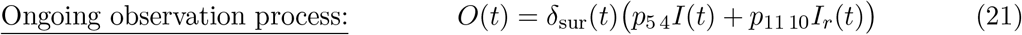

A number of the SCLAIV parameters can be independently estimated directly from etiological studies of the progression of disease in individuals under observation, treatment or hospitalization. These include the parameters *π*_lat_, *π*_asy_ and, *π*_rec_ (Table 5.1). The reduced infectivity *ε* of asymptomatic cases, though somewhat more difficult to obtain directly from case studies, may also be estimated independently from measurements at the rates at which asymptomatic individuals shed pathogens relative to symptomatic individuals.

The mortality rate parameter *α* can be estimated directly from mortality data. It appears to vary greatly from country to country in the case of Covid-19, ranging from *>* 10% for some Western European countries through 5-6% in the US to *<* 1% for some African and Middle Eastern countries (Worldometer Covid-19 Country Data Table). Of course, the reliability of these numbers depends on many factors, including surveillance levels, healthcare infrastructure, and willingness of the country to be transparent about its ongoing epidemic.

With respect to the two parameters *π*_thw_ and *π*_succ_, their ratio rather than their absolute values largely determines the proportion of individuals that return to S versus move onto L (cf. Eq. 18). Thus, as a first cut, we set *π*_thw_ = 1, to reduce by 1 the number of parameters that need to be estimated in the model. Further, as borne out in our fits to real data below, the parameters *κ* and *π*_succ_ are highly correlated. So, also as a first cut, it may be useful to also set *π*_succ_ = 1, in which case Eq. 19 implies that we are essentially estimating *β* as a fixed proportion of our *κ* estimate when the quarantine driver *δ*_qua_(*t*) is zero. This is an insight that we only verified by fitting both *κ* and *π*_succ_ to the data in the South African and English case studies discussed here. In future studies we will begin by setting *π*_succ_ = 1 as well and then evaluate the effects of releasing this constraint on the results we obtain.

### 3.3 Initial Conditions

It is uncertain how many infectious individuals are present in the population when the first case of a disease is identified in a particular population. Identification will be based on the presentation of symptoms, but will also depend on the the initial surveillance level *δ*_sur 0_, which cannot be estimated without considerable testing that only comes at a later stage in monitoring an outbreak. Thus the initial value *I*_0_ will need to be estimated. Its value will depend on the initial value that we assign to surveillance. In the absence of historic data that provides some estimate for *δ*_sur 0_, we might want to compare fits obtained for nominal assignments *δ*_sur 0_ = 0.1 and *δ*_sur 0_ = 0.5, noting that the smaller the value we select for *δ*_sur 0_, the concomitantly larger the estimates of *I*_0_ will be, when using Eq. 20 to fit the model to observed incidence.

If we want to reduce the number of parameters that need to be estimated, we can roughly base initial estimates of *A*_0_ and *L*_0_ on *I*_0_ as follows. If the average individual remains in *I*_0_ for a period *π*_rec_ and, if during this average time *π*_rec_ that it takes an individual to move through the I class the number of individuals in I has grown by an amount *G* to the level *GI*_0_ then we need to ensure that on average *G/π*_rec_ individuals move into the I class each unit of time during the period *π*_rec_. If each individual remains in A for an average period *π*_asy_, then we require

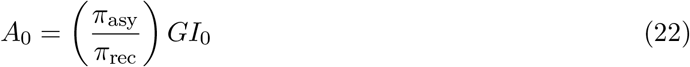

to ensure sufficient individuals move from A to meet the level of growth G. A similar argument of how many individuals are initially needed in disease class L to produce *A*_0_ leads to the equation

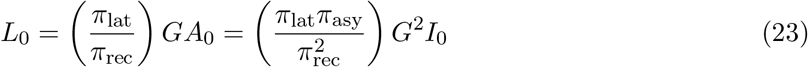

These estimates can always be checked to be reasonable, once the fit has been made and the initial outbreak simulated. This approximation, however, may be more useful for Bayesian Monte Carlo Markov chain (MCMC) [29, 30] than maximum likelihood (ML) estimation since since the former is much more computationally demanding than the latter.

The initial value *C*_0_ is more complicated because, beyond flows from C to A, it depends on flows back and forth between compartments S and C, as well as flows from C to CR when the quarantine driver is switched on. Thus, as with *I*_0_, we decided to include *C*_0_ as a member of the set of parameters whose values we would estimate by fitting model simulations to the initial set of incidence data.

When estimating parameters of an outbreak in an epidemiological naïve population, as is the case with an emerging disease such as Covid-19 (but generally not for measles or influence), it is reasonable to set *V*_0_ = 0. Even if this is not the case, whenever *V*_0_ is likely to be much smaller than *N*_nom_ then setting *V*_0_ = 0 will hardly affect the estimation process.

## 4 Parameter Estimation and Simulation Procedures

### 4.1 Estimation approaches

From the discussion in the previous, and prior to the onset of any drivers other than background surveillance and isolation/treatment rates, the set of parameters that we likely need to directly estimated from fitting our SCLAIV model to an initial Covid-19 incidence outbreak times series of say 30 or 40 points is

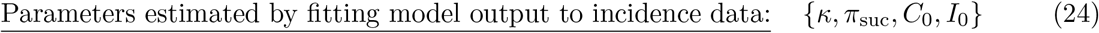

Once various drivers are turned on (e.g. social distancing, quarantining), or go into switching mode (e.g. surveillance), then additional parameters will need to be fitted as discussed in the context of our analysis of the England outbreak, as discussed below.

We used a maximum likelihood (ML) approach, as described in [15], to obtain our first best estimate of the parameters 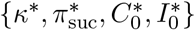. We then used these ML estimates (^*∗*^ values) to set the means of *a priori* distributions used to implement a Monte Carlo Markov chain Bayesian estimation procedure [29, 30], as described in more detail in the supplementary material.

Beyond our initial fit, we assumed that one or more of the drivers came into operation, either implicitly (e.g., social distancing) or explicitly (increase in surveillance and quarantine rates) at points in time after the start of the outbreak. The parameters defining these emerging implicit drivers could be estimated by fitting model output to incidence data streams beyond those used to fit the basic SCLAIV process, while the parameters associated with the implemented explicit drivers will come from information on the application of these drivers to the population.

When estimating these drivers from incidence data beyond the initial data used to estimate the basic SCLAIV parameters, as a first tack driver x for example could be regarded being constant once switched on. In this case, in Eq. 16 we set *δ*_x ∞_ = *δ*_x 0_, nominally set *σ*_x_ = 1 and *t*_x 1*/*2_ = 1000 say, and then estimate the two parameters *t*_x 0_ and *δ*_x 0_. If this does not provide a good enough fit, we can more general fit all five parameters {*t*_x 0_, *δ*_x 0_, *δ*_x ∞_, *t*_x 1*/*2_, *σ*_x_}that define the form depicted in Eq. 16, or all but one of the parameters if we set *σ*_x_ = 2 for a gradual switch, *σ*_x_ = 5 for an intermediate switch, or *σ*_x_ = 20 for an abrupt switc (Fig. 2).

### 4.2 Forecasting and Scenario Analyses

One can usefully identify three time periods of interest associated with parameter estimation, forecasting and policy scenario analyses for the population of interest: [0, *t*_est_], [*t*_est_, *t*_data_], and [*t*_data_, *t*_forecast_], where *t*_est_ ≤ *t*_data_ is the interval over with the parameter estimation procedure is conducted, *t*_data_ is the length of the data time series and *t*_forecast_ ≥ *t*_data_ is the end of the forecast interval.

Credibility interval projection and counter factual scenario analyses can be undertaken by generating an ensemble of stochastic runs on the interval [*t*_est_, *t*_data_] and either using randomly selected subsets of the parameter vectors (*κ, π*_suc_, *C*_0_, *I*_0_) that were accepted by an MCMC procedure (credibility interval projections) or using such subsets with the addition of selecting counterfactual driver parameter values to address “what if” questions such as, “what if social distancing had been implemented on a specified date?”

Forecasting studies can be undertaken by generating ensembles of stochastic runs on the interval [*t*_data_, *t*_forecast_], using randomly selected subsets of parameter pairs (*κ, π*_suc_) that were accepted by an MCMC procedure. We note that in these forecasts the ML or MCMC estimated parameters (*C*_0_, *I*_0_) are no longer relevant since the initial conditions for these stochastic simulations are the estimated state of system—e.g. from an ML optimization—at time *t*_data_. In addition, policy scenario forecasts can also be made by setting driver parameters to values reflecting the policy scenarios under consideration such as “What if surveillance or quarantine rates are increased?” or “What will be the effects of rolling out vaccination strategies at different specified rates?”

We undertook our ML optimizations and forecasting simulations using a Numerus Model Builder Data Analysis and Simulation (Covid-19 NMB-DASA) web app built for the purpose of carrying out disease outbreak studies using the SCLAIV+response model framework (a description of this web app and discussion of its use can be found in our supplementary material).

## 5 Covid-19 Case Study

The South African and English Covid-19 studies presented here are of limited scope. Studies that dig deeper than illustrated here require teams of individuals that include data professionals who are familiar with any idiosyncrasies in the data. Comprehensive forensic analyses of outbreak data require deeper insights into the data than can be obtained when downloading incidence and mortality time series from the web. Thus the brief studies presented below are meant to be purely illustrative rather than comprehensive.

### 5.1 Parameters for Generic Covid-19

In Table 5.1 the parameter values are listed for modeling the initial phase of non-specific Covid-19 outbreaks. Values specific to particular regions come from fitting an epidemic model to the initial incidence and mortality outbreak data. Response driver parameter values will, of course, be region specific and depend on policies implemented to help control local outbreaks.

### 5.2 Covid-19 in South Africa

The first case of Covid-19 was recorded in South Africa on March 5 and a week later only 1 of the 17 recorded cases appeared to involve a local transmission event (VOA News, March 12, 2020): most new cases during the first three weeks of March 2019 were associated with individuals returning from trips to Europe and the local outbreak only started to pick up the last week of March. This local importation effect is event in the new cases plotted in Fig 4A (see region around the last week of March). Thus we decided to only start fitting the new case data from March 28 onwards. We also fitted our model to a 7-day lagged moving average (Fig 4A), rather than the raw data itself. The reason for this is the strong seven-day oscillation evident in new case reported around the world [35], as well as the fact that newly identified cases are likely associated with individuals actually transferring to the I class some days prior to this identification event.

**Figure 4:**
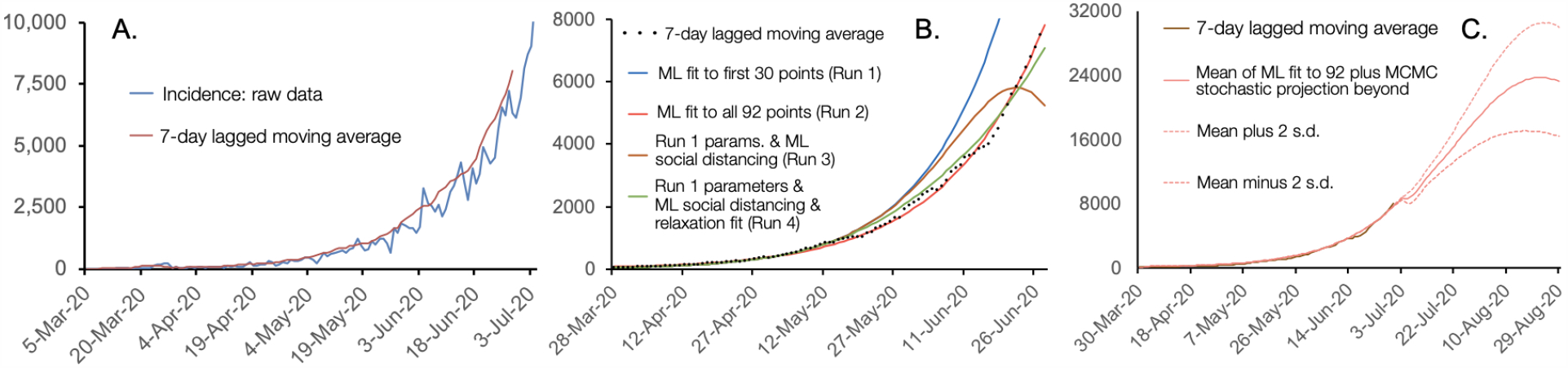
**A**. The incidence (blue) data time series and a 7-day lagged moving average (brown) plotted for new cases of Covid-19 in South Africa from March 5 to June 28 (raw data to July 4), 2020. **B**. Maximum-likelihood (ML) plots for fitting the 4 basic SCLAIV model parameters listed in Eq. 24 to the data from March 28 to April 27 (30 points) and extrapolated out to June 28 (Run 1: blue) and to the data from March 28 to June 28 (Run2: light red) with other parameters as listed in Tables 5.1 and 5.2. Runs 3 (brown) and 4 (green) fix the 4 basic SCLAIV parameters to the values estimated in Run 1, but Run 3 allows for an ML estimation of the five social distancing parameters (Table 5.2) and Run 4 for ML estimation of three of five social distancing and three of five social relaxation parameters (see Table 5.2 for details). **C**. Mean plus/minus 2 standard deviation projections of incidence starting at the estimated June 28 state and ending on August 29, using parameters *κ, π*_suc_, *I*_0_ and *C*_0_ selected from a posterior distribution obtained from a Monte Carlo Markov chain (MCMC) fit to the 7-day lagged moving average incidence data. Data source: Statista South African Covid-19 Study.

We first used maximum likelihood optimization [15] to fit our model to the first 30 points in our 7-day lagged moving average incidence data (Run 1, Fig 4B) and then compared this to fitting all 92 points of these data (Run 2, Fig 4B). Two additional runs at fitting these 92 incidence data points yielded fits that in one case (Run 3, Fig 4B) was almost as good as the first (Run 2) and another marginally better (Run 4, Fig 4B; error differences between fits varied by 1 or fewer parts per ten million—see Table 5.2).

Although maximum likelihood (ML) fits to the March 28-June 28 incidence data (92 points) look incredibly good (Run 2, Fig 4B), an ML fit to the first 30 points (March 28-April 27; Run 1, Fig 4B) showed significant divergence from the data around May 10 (Fig 4A; blue curve). This 30 point fit, however, can be brought back in line with the data from May 10 to mid-June (Fig 4A; blue curve), by switching on social distancing after day 30 at an essentially constant rate *δ*_sod_ = 0.005 (with contact rate reduction is *δ*_con_ = 0.1; Run 3, Fig 4B). During the last week of June, however, this new curve under-fits the data, possibly because the social distancing driver begins to be relaxed as individuals start to return to more normal behavior. We note that the projected impact of social distancing at the rate *δ*_sod_ = 0.005 over the period April 27 to June 28 (differences between Runs 1 and 4 in Fig 4B) is an estimated around 82 thousand surveilled cases averted (or an estimated 162 thousand actual cases when surveillance is at 50%, i.e., *δ*_sur_ = 0.5).

A Bayesian MCMC estimation procedure [30] was used to obtain a maximum-likelihood posterior distribution of the quadruple (*κ, π*_suc_, *I*_0_, *C*_0_) associated with fitting the model to the first 92 data points. A projection of the incidence beyond this point (i.e., June 28) until August 29 was made, as described in Section 4.2. These values were then used in 100 stochastic runs of the model to produce the credibility bounds provided by the mean ± 2 standard deviations graphed in Fig. 4C. We note that the slight dip in the mean projections at the start of the forecast are due to the fact that the range of parameter pairs (*κ, π*_suc_) obtained from our MCMC analyse varied across an order of magnitude, though the correlation between the values of these two parameters was exceptionally high (.0.99; see our supplementary material).

These MCMC projections suggest that incidence rates in South Africa will increase to between 15 and 30 thousand surveilled cases per day (half the number of actual cases given *δ*_sur_ = 0.5 in the model) by the end of August if stronger social distancing measures are not effected during July and August, 2020. This estimate, however, could well be conservative because the observed flatting of the incidence curve around mid August (Fig 4C) is due to reductions in the proportion of susceptibles (an artifact of setting *N*_nom_ to 10 million when the population of South Africa is close to 60 million) rather than the action of epidemic drivers. This effect would be much weaker if the nominal population size used in our simulations was several times larger. We stress, however, that this narrative does not account for effects that would arise from the spatial structure of the South African population. Accounting for spatial structure would, of course, require a much deeper analysis of the outbreak than can be investigated using any kind of homogeneous SEIR or SCLAIV model.

### 5.3 Covid-19 in England

The first two confirmed cases of Covid-19 in England identified on January 29 in the city of York (Metro News, April 19, 2020). During the month of March another 56 cases in England were recorded (UK Government Sources). Our analysis starts at this point in fitting our model to new case in England on March 1 to June 30, as computed in terms of a 7-day lagged moving average (i.e., incidence on the March 1 was on actual incidence averaged across the week of February 24 to March 1) to deal with the fact that incidence occurs on or before the day it is reported and a strong 7-day cycle has been observed in data collected around the world [35].

By including the ramping up of social distancing behavior in Fit 2 (Fig. 5, silver curve) to the first 60 data points (i.e., through March and April), we see that we capture the peak incidence that occurs around the middle of April, but the subsequent fall to near zero around the middle of June is too optimistic, presumably because of the relaxation of social distancing that occurs throughout May. By adding social relaxation to our estimation process in Fit 3 (Fig. 5, dark yellow curve) of all the data (March 1 to June 30), a much better fit is obtained for the months of May and June, but not for the initial months of March and April. Thus, in addition to estimating social distancing and social relaxation in Fit 4 (Fig. 5, blue curve) we also estimated changes in surveillance over time. We now obtained a much better fit (last four significant digits in the absolute log-likelihood value dropped from 9559 in Fig 3 to 1999 in Fit 4: see Table 5.3).

**Figure 5:**
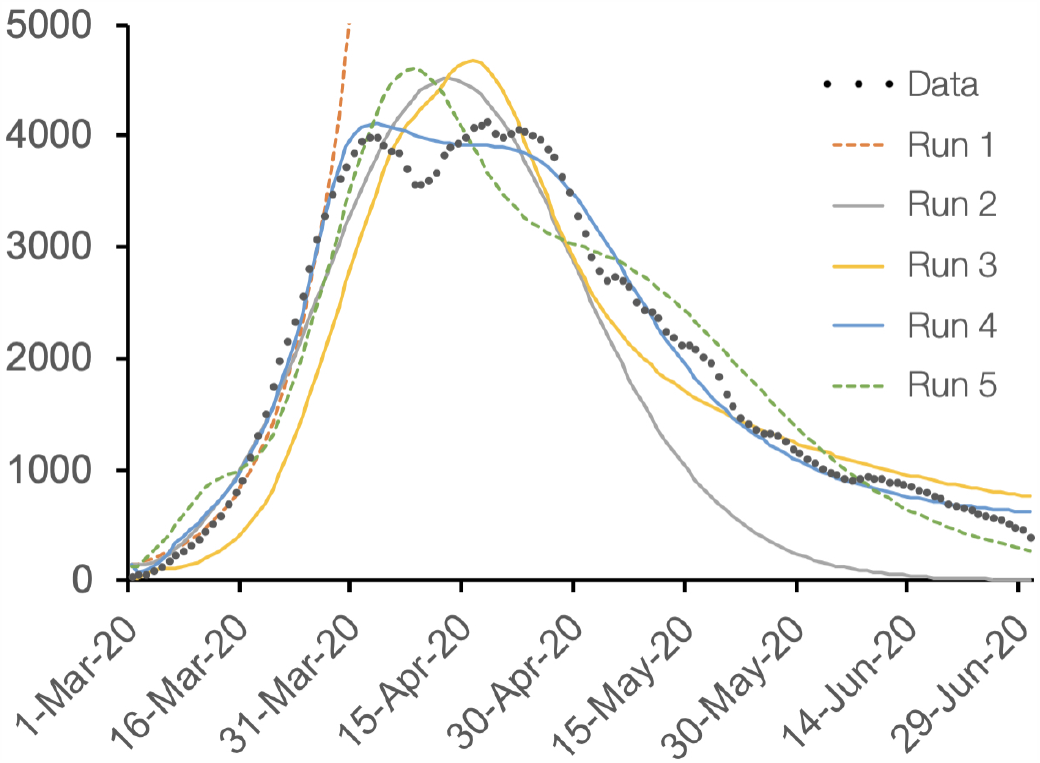
Five different fits (parameters reported in Table 5.3) to a 7-day lagged moving average of Covid-19 new case (incidence) in England (black dots) plotted for March 1 to June 30 (using raw data to July 6), 2020. Data source: UK Government Sources. Run 1 (broken orange—only initial growth phase shown) and 2 (silver) are fits of the four epidemic parameters listed in Eq. 24 alone to the first 30 points and all 122 points (including initial values) respectively. Runs 2 (silver), 3 (dark yellow), and 4 (blue) progressively include switching on social distancing, social relaxation and ramping up surveillance. Run 5 (broken green) is the same as Run 4 except now surveillance is ramped down rather than up.

Fit 4 implies that social distancing begins during day 21 at a rate of 0.18 per day with a switching point on day 41 and a final rate of 0.80 per day. It also implies that social relaxation begins on day 15 at a rate of 0.06 per day, with a switching point on day 129 and a final rate of 0.46 per day. The fact that social relaxation has an earlier onset than social distancing might seem odd, but it is not problematic. It arises because we did not constrain the onset of social distance to occur before the onset of social relaxation, but has no effect on projects because there are no individuals in class SR until the onset of social distancing. Thus its effect is the same as if the onset of social relaxation was also on day 21 and the initial social relaxation rate were 0.12=0.18-0.06—calculations correct to 2dp). Fit 4 also implies an increase in surveillance from 26% on March 1 with a switching point 5 days later and a maximum surveillance level of 67%.

For purposes of comparison, in Fit 5 (Fig. 5, broken green curve) we estimated the same set of parameters as in Fit 4, except now we constrained the final surveillance value to be less than the initial surveillance value, as might conceivable (though unlikely) occur if a healthcare surveillance system broke down during the course of a severe outbreak. In this case a reasonable fit was still obtained, with Fit 5 being a better fit than Fit 3 (9559 vs 6008 for the last four significant digits in the absolute log-likelihood value: see Table 5.3). The epidemics represented by these two incidence curves, however, are vastly different. In Run 4, the estimated percent of individuals that are immune (*V*) on June 30 is 7.6% while in Run 5 it is 87.7%. Run 4 is much more credible than Run 5 because we have no evidence that the English surveillance process has broken down over time. Our comparison of Runs 4 and 5 highlights that without additional data, such as the proportion of individuals in the population that are sero-positive, any estimation process is not securely anchored in reality and model estimation results maybe very misleading.

In the five fits described above, we did not fit the disease induced mortality rate parameter *α* to mortality data, although a constant for this value can be directly estimated by looking at the number of deaths to the total number of cases that have been observed so far. Constant values of *α*, however, are unlikely because the healthcare sector becomes better at identifying and treating cases over time. A disturbing set of statistics, however, is the range of mortality rates across different countries that, as previously mentioned, range from *<* 1% to *>* 10%. This leads one to believe that either incidence or mortality have either been deliberately or inadvertently under reported in many countries, or the Covid-19 really does express itself differently in countries are environmentally vary different (e.g., climate, density, or socioeconomic factors).

## 6 Discussion and Conclusion

In generalizing the SEIR framework for modeling disease outbreaks to a SCLAIV+D setting, we have allowed for the “exposed class” (E) to reflect the reality that not all exposed individuals become infected, a situation that is applicable to all disease transmission processes. In our particular exposition of fitting the model to Covid-19 outbreaks in England and South Africa, we did not consider adding a quarantine process. Effective real-time contact tracing and consequent quarantining requires the implementation of digital methods that are faster, more efficient, and more scalable than are currently available [36], though limited efforts in Iceland and Singapore appear to have met with some success [37, 38]. Getting an independent fix through contact tracing on the contact rate parameter *κ* is important, however, since in our MCMC study of the South African data, *κ* and the succumbing period *π*_suc_ are highly correlated (supplementary material). Thus, fixing one of these two parameters ahead of time makes the estimation process more stable and efficient. Of course, in the absence of intensive transmission studies, such as [16], there is an element of subjectivity to measuring *κ*. For sexually transmitted disease, the definition of a contact is quite clear. But for disease spread through aerosols and fomites, contacts require thresholds for distance, duration, and environmental context for putative contact events to be scored as a pathogen transmission event [39] that the individual can either thwart or succumb to through physical barriers or an innate immune response.

The SCLAIV setting also includes both asymptomatic and symptomatic pathogen transmitting individuals, where the asymptomatic class A includes both presymptomatic (progressing from A to I) and perfectly asymptomatic (progressing from A to V) individuals. The assumption we made that the rates at which individuals in both A and I recover (i.e., the rates of progression from A to V and I to V) are the same was purely for convenience (i.e., to reduce parameters from proliferating in the model). Clearly this assumption can be removed and a recovery rate of asymptomatic individuals that do not progress to symptomatic can be reliably estimated directly from data once better individual testing and monitoring protocols become available. We may have to wait for the next Covid-like outbreak to occur for such protocols to be in place, as well real-time contact tracing and quarantining protocols and technology, in response to lessons learned from the current Covid-19 pandemic.

Our SCLAIV model can be embedded in a more general population demographic (age and sex, as well as healthcare worker and other high risk categories) and spatial structure setting, as depicted in Fig. 1. Again, the kind of data needed to support the additional parameters that would justify the complexities involved in such an embedding are not now available, although some age and sex-related Covid-19 transmission and mortality data have been published [40, 41]. Most models that include spatial structure, for example, when carrying out analyses using real data have very simply underlying epidemiological descriptions [42]. If spatial structure is not included when considerable levels of spatial heterogeneity exist, then incidence curves tend to broaden [13] and multiple peaks, such as that observed in the England data (Fig. 5) may occur. Such curves when fitted to homogeneous SEIR or SCLAIV model underestimate the epidemics reproductive potential (as measure by *R*_0_) [13], and a metapopulation model setting [15, 26] is needed to obtain accurate results.

A clearer understanding of how epidemics respond to the implementation of drivers in rolling out policies to “flatten the (incidence/hospitalization) curve” is essential to managing outbreaks so that healthcare facilities do not become overwhelmed. Thus, considerable interest exists in identifying, or in inferring from the data [30], the times when driving processes (increased surveillance, social distancing and subsequent relaxation, contact tracing with quarantining, rolling out vaccination programs when available) have either emerged through behavioral changes or should be applied as policy measures. Procedures for statistically identifying driver onset points have been developed in the context of an SIR model [30]. Our SCLAIV+response formulation presented here represents the first time a model that explicitly includes driver dynamics has been forensically applied to identifying driver onset and evaluating driver effects in any disease outbreak data.

Ultimately, however, as our fitting of the England Covid-19 data illustrate, all fitting procedures should be treated with extreme caution unless the fits can be anchored using additional data. The most informative of these are likely to be serology data that provide an accurate assessment of the size of the V class at one or more points in time. Of course, not all serology tests are accurate [43], so we should do all we can to, at least, evaluate the sensitivity of any tests that are implemented so that accuracy estimates of the size of the immune class are possible. In short, we should not underestimate the value of being able to measure immunity levels at points in time when models are being used to devise the most appropriate policies for managing outbreaks. For this reason, it is not worth pursuing a more in depth analysis of the South Africa, English, or any other country for that matter than undertaken here, without access to additional data that can only come from working with healthcare authorities responsible for collecting and curating outbreak and other related data for any region for which models are being used to formulate policy.

Finally, Covid-19 is but one member of the class of diseases for which our SCLAIV+response formulation is appropriate. For reasons relating to our inability to collect appropriate data, it has not made sense in the past to explicitly divide the exposed class in SEIR models into separate contact and latent-infection classes. With the advent of appropriate technologies in the future to implement real time contact tracing and subsequent quarantining [36], our SCLAIV+response elaboration of the SEIR model should provide a step forward in improving the adequacy of dynamics models [44] in formulating policy and forecasting disease outbreaks.

**Table 1:**
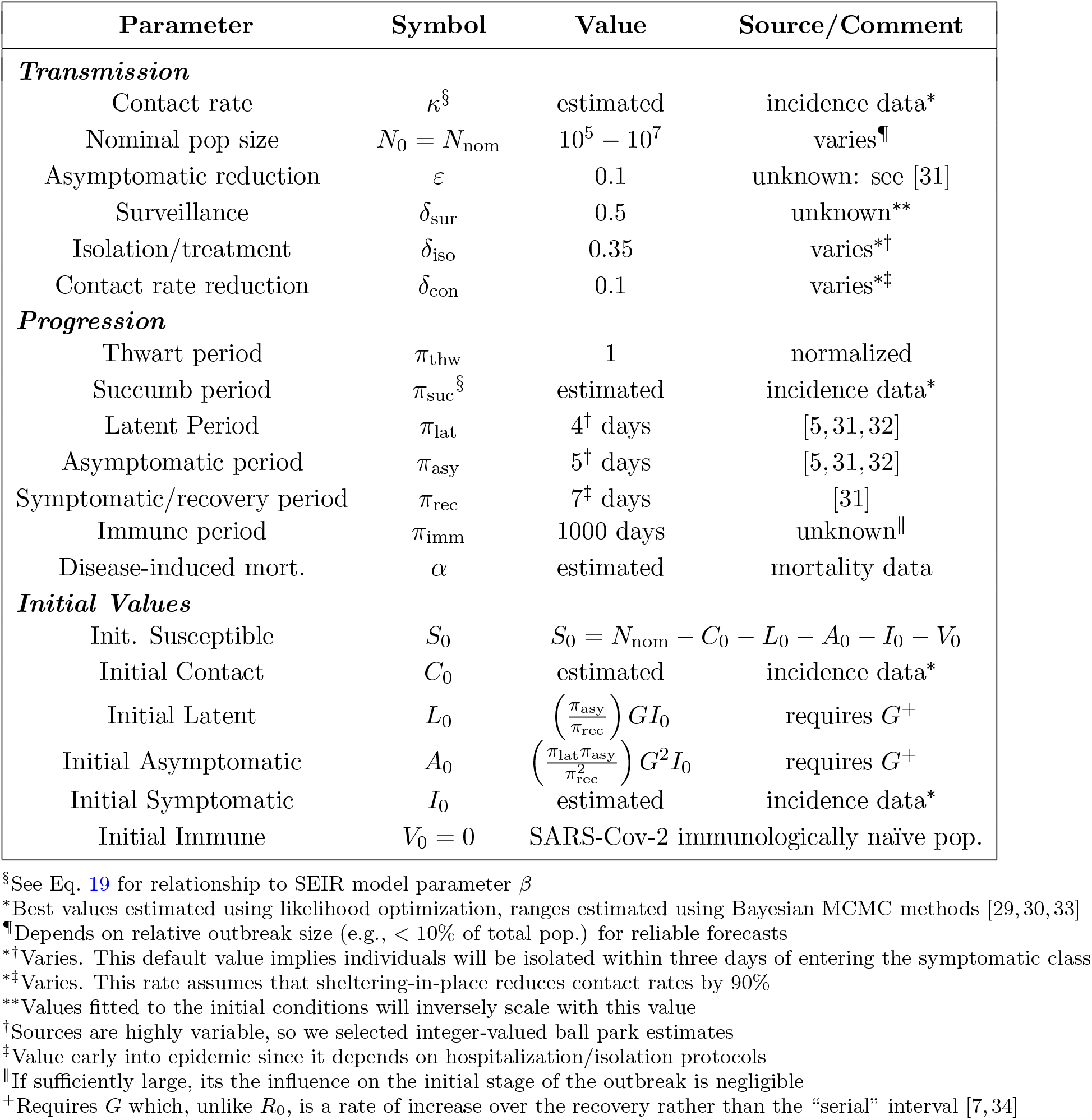
Parameters for fitting and simulating a basic Covid-19 SCLAIV outbreak on [0, *t*_est_])

| Parameter | Symbol | Value | Source/Comment |
| --- | --- | --- | --- |
| **Transmission** |  |  |  |
| Contact rate | $\kappa^{\S}$ | estimated | incidence data* |
| Nominal pop size | $N_0 = N_{\text{nom}}$ | $10^5 - 10^7$ | varies<sup>¶</sup> |
| Asymptomatic reduction | $\varepsilon$ | 0.1 | unknown: see [31] |
| Surveillance | $\delta_{\text{sur}}$ | 0.5 | unknown** |
| Isolation/treatment | $\delta_{\text{iso}}$ | 0.35 | varies*<sup>†</sup> |
| Contact rate reduction | $\delta_{\text{con}}$ | 0.1 | varies*<sup>‡</sup> |
| **Progression** |  |  |  |
| Thwart period | $\pi_{\text{thw}}$ | 1 | normalized |
| Succumb period | $\pi_{\text{suc}}^{\S}$ | estimated | incidence data* |
| Latent Period | $\pi_{\text{lat}}$ | 4<sup>†</sup> days | [5, 31, 32] |
| Asymptomatic period | $\pi_{\text{asy}}$ | 5<sup>†</sup> days | [5, 31, 32] |
| Symptomatic/recovery period | $\pi_{\text{rec}}$ | 7<sup>‡</sup> days | [31] |
| Immune period | $\pi_{\text{imm}}$ | 1000 days | unknown |
| Disease-induced mort. | $\alpha$ | estimated | mortality data |
| **Initial Values** |  |  |  |
| Init. Susceptible | $S_0$ | $S_0 = N_{\text{nom}} - C_0 - L_0 - A_0 - I_0 - V_0$ | |
| Initial Contact | $C_0$ | estimated | incidence data* |
| Initial Latent | $L_0$ | $\left(\frac{\pi_{\text{asy}}}{\pi_{\text{rec}}}\right) G I_0$ | requires $G^+$ |
| Initial Asymptomatic | $A_0$ | $\left(\frac{\pi_{\text{lat}} \pi_{\text{asy}}}{\pi_{\text{rec}}^2}\right) G^2 I_0$ | requires $G^+$ |
| Initial Symptomatic | $I_0$ | estimated | incidence data* |
| Initial Immune | $V_0 = 0$ | SARS-Cov-2 immunologically naïve pop. | |

<sup>§</sup>See Eq. 19 for relationship to SEIR model parameter $\beta$
\*Best values estimated using likelihood optimization, ranges estimated using Bayesian MCMC methods [29, 30, 33]
<sup>¶</sup>Depends on relative outbreak size (e.g., < 10% of total pop.) for reliable forecasts
\*<sup>†</sup>Varies. This default value implies individuals will be isolated within three days of entering the symptomatic class
\*<sup>‡</sup>Varies. This rate assumes that sheltering-in-place reduces contact rates by 90%
\*\*Values fitted to the initial conditions will inversely scale with this value
<sup>†</sup>Sources are highly variable, so we selected integer-valued ball park estimates
<sup>‡</sup>Value early into epidemic since it depends on hospitalization/isolation protocols
<sup>||</sup>If sufficiently large, its the influence on the initial stage of the outbreak is negligible
<sup>+</sup>Requires $G$ which, unlike $R_0$ , is a rate of increase over the recovery rather than the “serial” interval [7, 34]

**Table 2:**
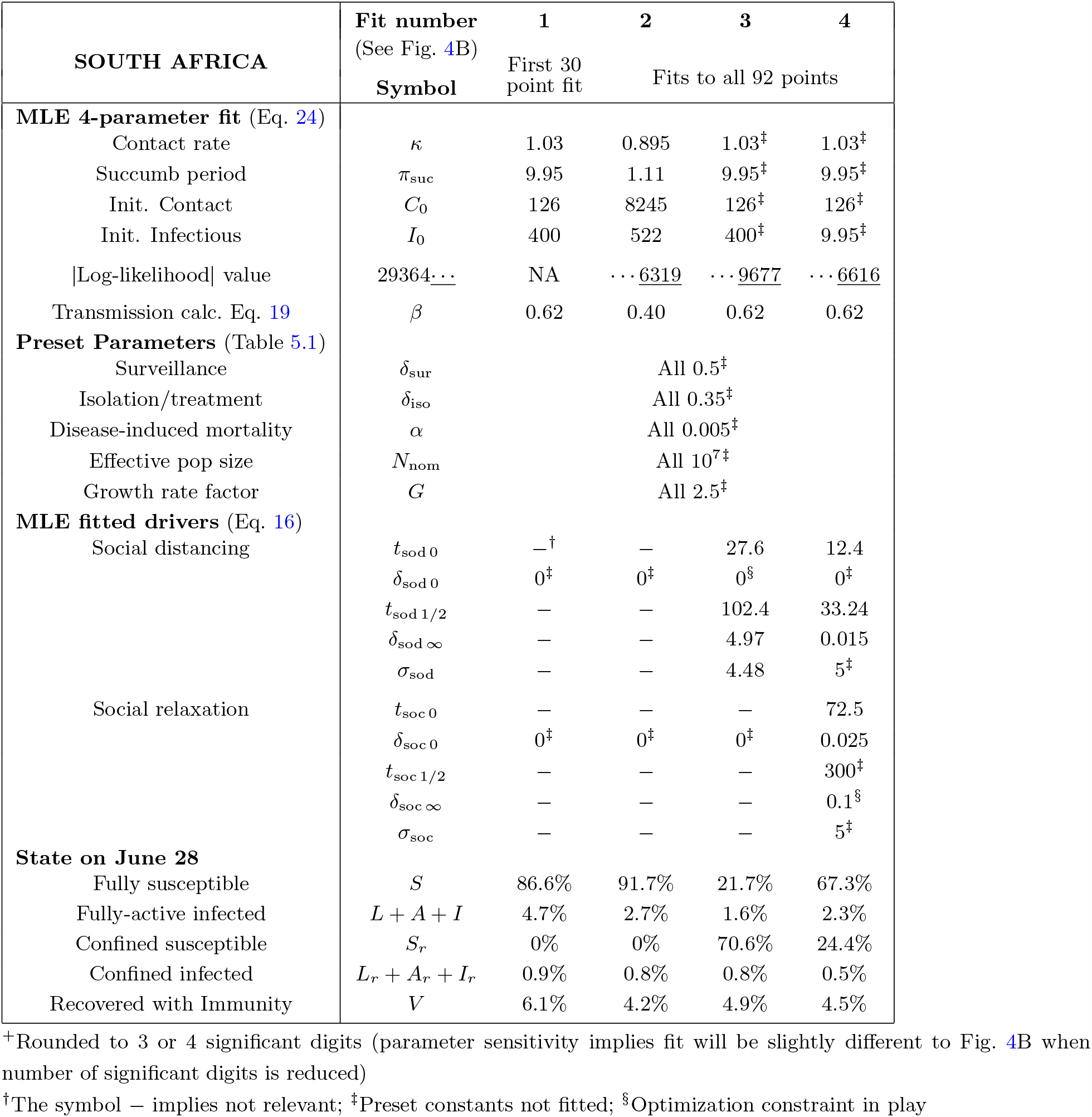
Specific values set and maximum-likelihood values obtained in fitting the South African Covid-19 incidence data (7-day lagged moving average) for March 28-June 30 (Fig. 4B). See Table 5.1 for general Covid-19 epidemic parameter values.

**Table 3:** Specific values set and maximum-likelihood values obtained in fitting the England Covid-19 incidence data on March 1 to June 30 (see Table 5.1 for other parameter values)

| ENGLAND, UK | Fit number | 1 | 2 | 3 | 4 | 5 |
| --- | --- | --- | --- | --- | --- | --- |
|  | (See Fig. 5)<br>Symbol | First 30<br>point fit | First 60<br>point fit | Fit to all points |  |  |
| **MLE 4-parameter fit<sup>+</sup>** (Eq. 24) |  |  |  |  |  |  |
| Contact rate | $\kappa$ | 1.71 | 1.71 | 2.85 | 2.19 | 2.41 |
| Succumb period | $\pi_{\text{suc}}$ | 0.194 | 0.194 | 0.947 | 1.502 | 0.062 |
| Init. Contact | $C_0$ | 1<sup>§</sup> | 1<sup>§</sup> | 5525 | 7620 | 4721 |
| Init. Infectious | $I_0$ | 959 | 958 | 103 | 1238 | 873 |
| \|Log-likelihood\| value | 35703... | NA | NA | ...9559 | ...1999 | ...6008 |
| Transmission Eq. 19 | $\beta$ | 0.28 | 0.28 | 1.21 | 1.07 | 0.14 |
| **Preset Parameters** (Table 5.1) |  |  |  |  |  |  |
| Isolation/treatment | $\delta_{\text{iso}}$ | | | All fits 0.35 | | |
| Disease-induced mortality | $\alpha$ | | | All fits 0.01 | | |
| Effective pop size | $N_{\text{nom}}$ | | | All fits $10^7$ | | |
| Growth rate factor | $G$ | | | All fits 2.5 | | |
| **MLE fitted drivers** (Eq. 16) |  |  |  |  |  |  |
| Surveillance | $t_{\text{sur } 0}$ | —<sup>†</sup> | — | — | 0<sup>‡</sup> | 0<sup>‡</sup> |
| | $\delta_{\text{sur } 0}$ | 0.5<sup>‡</sup> | 0.5<sup>‡</sup> | 0.5<sup>‡</sup> | 0.263 | 0.604 |
| | $t_{\text{sur } 1/2}$ | — | — | — | 4.6 | 11.5 |
| | $\delta_{\text{sur } \infty}$ | — | — | — | 0.671 | 0.056 |
| | $\sigma_{\text{sur}}$ | — | — | — | 5<sup>‡</sup> | 5<sup>‡</sup> |
| Social distancing | $t_{\text{sod } 0}$ | — | 5.2 | 16.5 | 21.6 | 17.0 |
| | $\delta_{\text{sod } 0}$ | 0<sup>‡</sup> | 0.051 | 0.074 | 0.177 | 0.096 |
| | $t_{\text{sod } 1/2}$ | — | 99.9 | 30.5 | 41.8 | 36.7 |
| | $\delta_{\text{sod } \infty}$ | — | 0.021 | 0.790 | 0.795 | 0.553 |
| | $\sigma_{\text{sod}}$ | — | 5<sup>‡</sup> | 5<sup>‡</sup> | 5<sup>‡</sup> | 5<sup>‡</sup> |
| Social relaxation | $t_{\text{soc } 0}$ | — | — | 33.5 | 15.5 | 11.5 |
| | $\delta_{\text{soc } 0}$ | 0<sup>‡</sup> | 0<sup>‡</sup> | 0.040 | 0.060 | 0.017 |
| | $t_{\text{soc } 1/2}$ | — | — | 150.8 | 129.6 | 59.5 |
| | $\delta_{\text{soc } \infty}$ | — | — | 0.202 | 0.455 | 0.342 |
| | $\sigma_{\text{soc}}$ | — | — | 5<sup>‡</sup> | 5<sup>‡</sup> | 5<sup>‡</sup> |
| **State on June 30** |  |  |  |  |  |  |
| Fully susceptible | $S$ | 4.2% | 0.1% | 6.8% | 19.2% | 4.4% |
| Fully-active infected | $L + A + I$ | 0.1% | 0.0% | 0.3% | 0.2% | 0.6% |
| Confined susceptible | $S_r$ | 0% | 87.6% | 79.3% | 72.7% | 6.5% |
| Confined infected | $L_r + A_r + I_r$ | 0.2% | 0.3% | 0.6% | 0.2% | 0.6% |
| Recovered with Immunity | $V$ | 95.5% | 11.9% | 12.9% | 7.6% | 87.7% |

<sup>+</sup>Rounded to 3 or 4 sign. digits (parameter sensitivity implies fit slightly different to Fig. 5)<sup>†</sup>The symbol “—” implies not relevant; <sup>‡</sup>Preset constants not fitted; <sup>§</sup>Optimization hit constraint

## Data Accessibility

All data used in fitting the models are accessible at the referenced sites.

## Data Availability

All data used in fitting the models are accessible at the referenced sites.

## Authors Contributions

WMG conceived the study, carried out the analyses apart from the MCMC fitting of the South African data, generated the final figures and tables, wrote the first draft of the manuscript. RS designed and coded the Numerus Model Builder (NMB) applications program and the NMB-DASA platform with user design feature input from WMG. LLV and RS built the NMB model for the MCMC analysis for a final MCMC analysis carried out by LLV along with report of the MCMC methods in the supplementary material. All authors provided edits to the final version of the manuscript.

## A Appendices

### A.1 Continuous-time, Donor-controlled Systems

Continuous-time, donor-controlled systems of equations can be compactly expressed in terms of a vectors of system’s variables **x** = (*x*_1_, …, *x*_*n*_)^*′*^ (^*′*^ denotes vector transpose), and a matrix *F* of elements *f*_*ij*_, *i, j* = 1, …, *n* by the equation

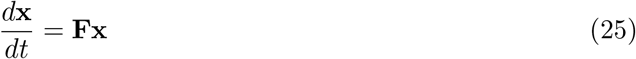

where the elements *f*_*ij*_, *i, j* = 1, …, *n*, expressed in terms of the donor controlled per-capita flow rates *γ*_*ij*_, (i.e. flows out of compartment *i* into compartment *j*),

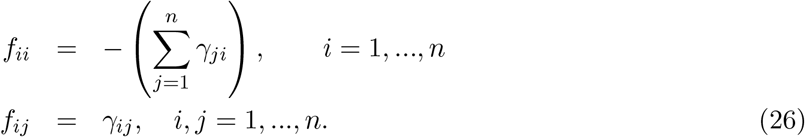

Under the assumption that no intrinsic growth or decay process occur within any of the disease classes (i.e., no births, non-disease induced mortality, or migration processes are included), it follows that *γ*_*ii*_ = 0, *i* = 1, …*n*. In this case, the equation for the *i*^*th*^ variable in systems Eq. 26 is

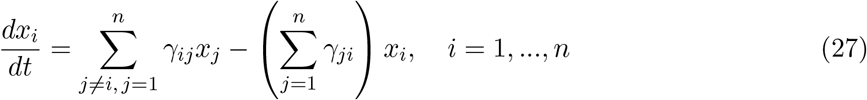

If all the rates in the system flow matrix Γ, with *ij*^*th*^ element

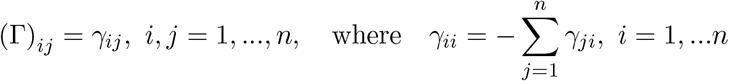

are constant or time dependent then the system is linear. Nonlinearities enter when one or more of the rates are dependent on the values of the various systems variable. Whether completely linear or not, we note that this system is subject to conservation principle invoked by the relationship 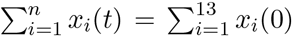 for all *t* because under the “no intrinsic growth or decay process” assumption it follows that

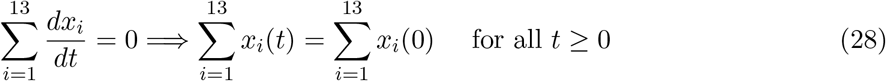

